# Intraoperative Plasma Proteomic Changes in Cardiac Surgery: In Search of Biomarkers of Post-operative Delirium

**DOI:** 10.1101/2022.06.08.22276153

**Authors:** Kwame Wiredu, Sean O’Connor, Erika Monteith, Brooke Brauer, Arminja N. Kettenbach, Hildreth R. Frost, Shahzad Shaefi, Scott A. Gerber

**Affiliations:** Department of Molecular and Systems Biology, Geisel School of Medicine at Dartmouth, Lebanon, NH; Program in Quantitative Biomedical Sciences, Geisel School of Medicine at Dartmouth, Lebanon, NH; Department of Anesthesia, Critical Care and Pain Medicine, Beth Israel Deaconess Medical Center, Boston MA; Department of Biochemistry and Cell Biology, Geisel School of Medicine at Dartmouth, Lebanon, NH; Dartmouth Cancer Center, Geisel School of Medicine at Dartmouth, Lebanon, NH; Department of Biomedical Data Science, Geisel School of Medicine at Dartmouth, Lebanon, NH; Department of Anesthesia, Critical Care and Pain Medicine, Harvard Medical School, Boston MA

**Author notes:** Corresponding author: Scott A. Gerber, Professor of Molecular and Systems Biology, 1 Medical Drive, Lebanon NH 03756.

**Keywords:** biomarker discovery, delirium, mass spectrometry, plasma, proteomics

## Abstract

**Purpose:** Delirium presents a significant healthcare burden. It complicates post-operative care in up to 50% of cardiac surgical patients with worse hospital outcomes, longer hospital stays and higher overall cost of care. Moreover, the nature of delirium following cardiac surgery with cardiopulmonary bypass (CPB) remains unclear, the underlying pathobiology is poorly understood, status quo diagnostic methods are subjective, and diagnostic biomarkers are currently lacking.

**Objective:** To identify diagnostic biomarkers of delirium and for insights into possible neuronal pathomechanisms.

**Experimental design:** Comparative proteomic analyses were performed on plasma samples from a nested matched cohort of patients who underwent cardiac surgery on CPB. A targeted proteomics strategy was used for validation in an independent set of samples. Biomarkers were assessed for biological functions and diagnostic accuracy.

**Results:** 47% of subjects demonstrated delirium. Of 3803 total proteins identified and quantified from patient plasma samples by multiplexed quantitative proteomics, 16 were identified as signatures of exposure to CPB, and 11 biomarkers distinguished delirium cases from non-cases (AuROC = 93%). Notable among these biomarkers are C-reactive protein, serum amyloid A-1 and cathepsin-B.

**Conclusions and clinical relevance:** The interplay of systemic and central inflammatory markers shed new light on delirium pathogenesis. This work suggests that accurate identification of cases may be achievable using a panel of biomarkers.

**Statement of Clinical Relevance:** The acute implication of delirium is well-documented, yet the true extent of the consequences beyond the immediate post-operative period has yet to be fully known. Despite its impact on the geriatric population, delirium remains underdiagnosed. Correctly identifying cases remain a challenge in clinical practice: the arbitrary and subjective nature of current diagnostic tools, such as the confusion assessment method, underscores the urgent need for diagnostic biomarkers. The clinical usefulness of delirium biomarkers extent beyond the objective identification of cases. Delirium biomarkers will also be useful for risk stratification, long-term follow-up of patients and may offer insights into possible etiologies that underpin the condition. In this report, we found systemic markers of inflammation with well-established association with delirium, as well as new biomarkers that shed new light on the condition. Although validation in a larger cohort is the necessary next step, our efforts lay the groundwork for future studies and highlight new frontiers in delirium research yet to be explored.

## Introduction

Delirium remains under-diagnosed in clinical practice[1–3]. Characterized by acute fluctuations in consciousness, deficits in attention and impairments in cognition not explained by a pre-existing neurocognitive disorder, delirium is etiologically heterogenous with a particularly high incidence after cardiac surgery[4, 5]. Following cardiac surgery, it complicates post-operative care in up to 50% of patients with increased length of hospitalization, increased mortality and higher overall cost of care[6]. In the long term, post-cardiotomy delirium patients are at increased risk of many complications, including re-admissions [7], cognitive decline [8–11], functional impairments [12], and stroke [13, 14], to mention a few. Clearly, delirium presents a significant healthcare burden on society. The true extent of the consequences beyond the immediate post-operative period remains unknown. Thus, the accurate identification of subjects for optimal care in the immediate post-operative period and for long-term follow-up is likely to exert a significant positive impact on patient care and costs if implemented successfully.

Unfortunately, many patients with delirium are missed [15, 16], an observation that is partly due to the subjective and variable nature of the current diagnostic approach. Efforts to improve recognition and accurate case identification has seen a steady rise in recent years, although a small fraction of these attempts has focused on biomarker discovery. Most of these biomarker studies also employed targeted quantification strategies for a sub selected list of genes or proteins, an approach that is inherently biased and blinded to potentially novel factors involved in the etiology or consequences of delirium [17][manuscript].

Challenges with delirium biomarker discovery are due, in part, to the lack of clarity regarding the underlying pathophysiology of the condition. While a one-size-fits-all explanation of delirium may be oversimplified, neuroinflammation induced by system-wide activation of an inflammatory cascade remains the prevailing mechanistic hypothesis[18, 19]. This is supported by recent untargeted and semi-targeted approaches that sought to study the proteome of human biofluids[20–27], although neuroendocrine and circadian dysregulation have also been reported[18]. The emerging focus on signaling and inflammatory markers necessitate biomarker discovery approaches that focus on the low-abundance proteome, using analytical platforms with the multiplexing capability and the requisite sensitivity to detect small changes in proteomic signatures.

In the present work, we comprehensively profiled the plasma proteome of subjects at baseline and post-cardiotomy for an untargeted analysis of the plasma proteome. We included abundant protein immunodepletion and peptide fractionation to enhance signal from the low abundance plasma proteome. Using independent set of samples, we validated candidate biomarkers at three time points (at baseline, post-bypass and post-operative) in order to understand the changing trajectories of these biomarkers over time as they relate to case identification. Finally, we demonstrate the diagnostic potential of a panel of candidate biomarkers, the accuracy of their use in discriminating cases from non-cases and the temporal association between intra-operative events and changes in biomarker levels.

## Results

### Clinical Profile of Study Participants

Subjects (*n* = 15) were selected from the parent study[28], which was a parallel group randomized controlled trial that enrolled 100 patients at Beth Israel Deaconess Medical Center (BIDMC), between July 2015 and July 2017. Delirium cases and non-delirium controls were age- and sex-matched (**Table 1**). There was no difference in baseline neurocognition between cases and non-cases, and the proportion of patients who received hyperoxic intraoperative treatment was comparable. There were no significant differences with regards to demographics, medical co-morbidities, pre-operative medications, or surgical characteristics. Details of the clinical characteristics of study subjects were reported previously[28].

**Table 1.** Selected baseline characteristics of study subject in the discovery phase. Delirium cases were age- and sex-matched to non-delirium controls. Details of all clinical characteristics of study subjects are reported elsewhere (Shaefi et al., 2021). Abbreviations: *t*MOCA: telephone-based Montreal Cognitive Assessment test for Dementia

### Discovery Phase of Biomarker Workflow

Using a multiplexed isobaric tagging (TMT)-based design, plasma samples at baseline and on post-operative day 1 from 7 delirium cases (CAM+) and 8 non-delirium controls (CAM-) were comprehensively profiled (**Figure 1)**. For precision, samples selected for the discovery phase of the study were analyzed in duplicates, for a total *n* = 60 samples, which necessitated the analysis of seven separate, batched multiplexes. To control for technical variation between batches, two channels in each of the seven 11-plex TMT sets were reserved as bridge samples using equal amounts of a pooled plasma sample. We fractionated the TMT-labeled peptides using off-line HPLC on a pentafluorophenyl (PFP) column as described previously[29] into 48 fractions, which were subsequently concatenated into 12 and analyzed by LC-MS/MS on an Orbitrap Fusion Lumos Tribrid instrument platform.

**Figure 1:**
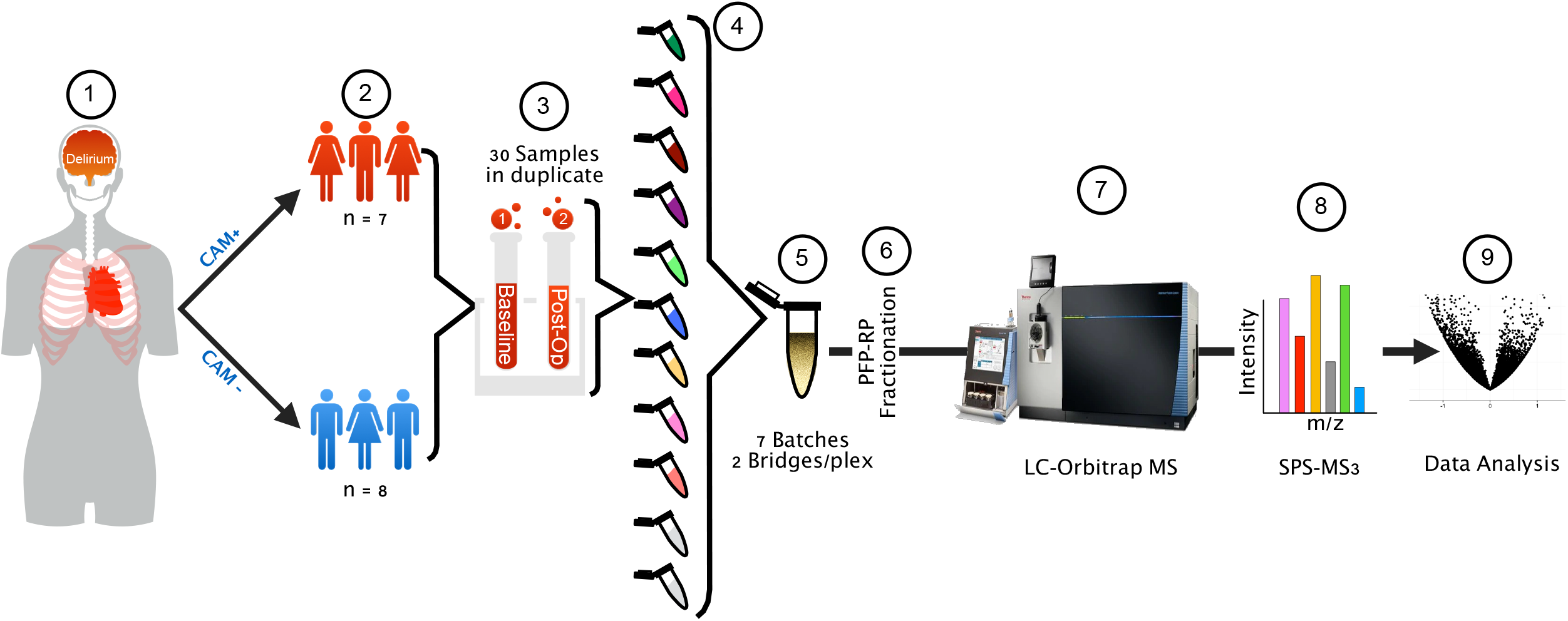
Study Design and Biomarker Discovery Workflow. Biomarker discovery: a cohort of 15 subjects were selected from the parent study of 100 patients who underwent a non-emergent coronary artery bypass grafting (CABG) on cardio-pulmonary bypass (CPB) as part of a previously published clinical trial (1,2). Plasma samples of delirium cases (CAM+) and non-delirium controls (CAM-) were retrieved from the biorepository for subsequent proteomic analysis (3). Samples were immunodepleted, digested and labeled with multiplex isobaric quantification (TMT) reagents. For each set of TMT reagents, two channels were reserved for bridge samples for post-hoc batch correction (4). TMT-labeled samples were concatenated (5) and additionally fractionated (6) prior to LC-MS/MS (7) for quantification at MS3 (8). After peptide spectral matching and false discovery rate (FDR) curation, the final dataset of 3803 proteins was quantified and analyzed for candidate biomarkers (9).

A collective total of 17,540 unique peptides from 3,803 proteins were identified from all seven multiplexes. An analysis of the number of proteins from each batch, separated into a binary group based on the corresponding number of peptides used in the identification of these proteins, demonstrates that our data are clearly dominated by so-called “one-hit proteins,” or proteins identified by a single peptide (**Figure 2A**). Often, single-peptide protein identifications are excluded from downstream analysis due to the increased risk of false protein identifications associated with single-peptide protein assignments. However, excluding all one-hit proteins can be a huge informational cost as some of these proteins may be biomarkers of interest.

**Figure 2:**
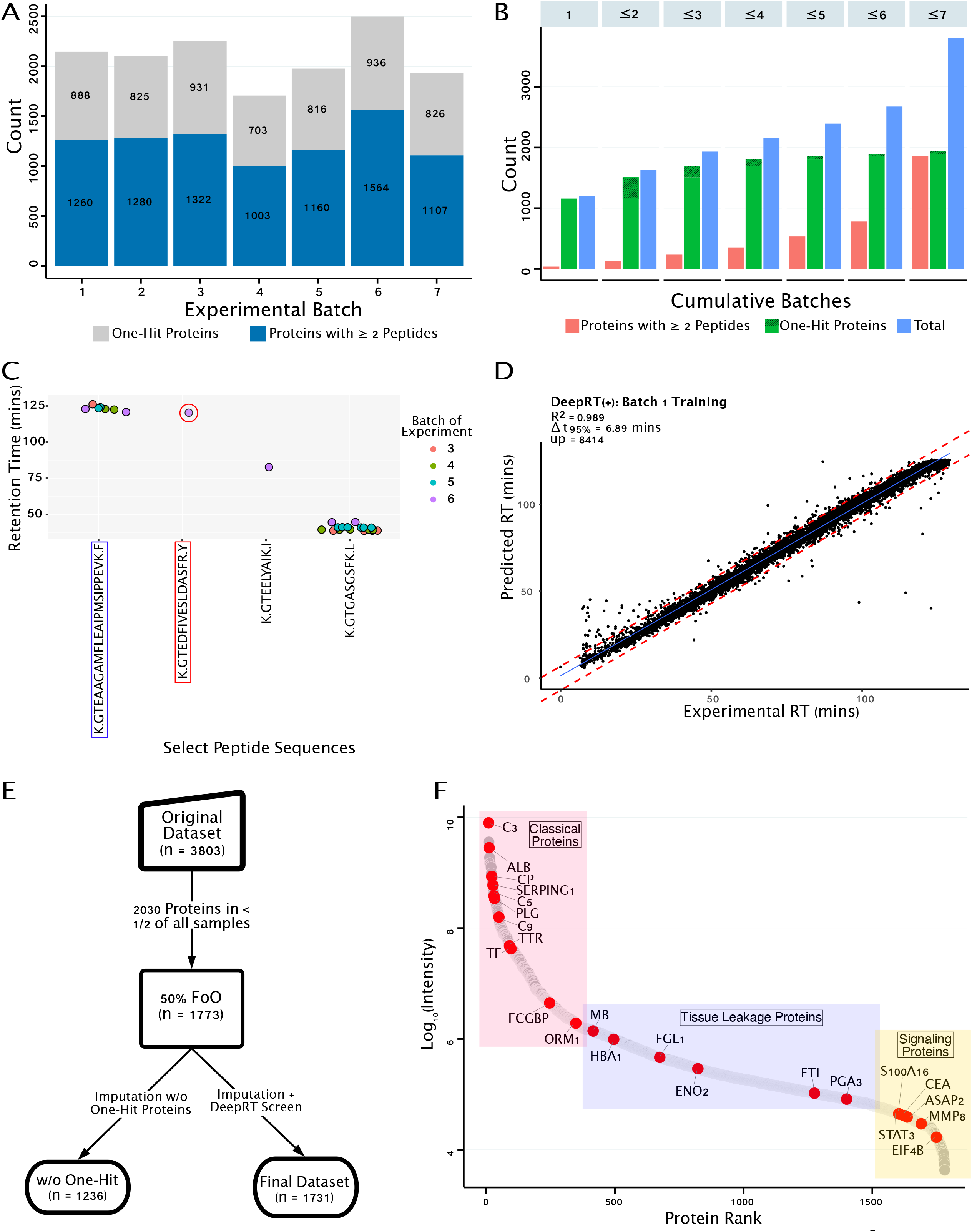
**A.** Total number of proteins identified per batch. Bars are demarcated by the number of unique peptides used for protein identification. Gray portion of each barchart represents proteins identified by only a single peptide, highlighting the scope of one-hit proteins in our analysis. **B.** Number of proteins identified in a cumulative number of experimental batches. For example, of the 1638 total proteins identified in up to two cumulative batches of experiments (cumulative batch ≤ 2), about 90% of those (*n* = 1470) were one-hit proteins. The number of one-hit proteins increases only marginally with increasing cumulative batches (light green portion of the green bars), in contrast to proteins identified from at least two peptides. **C.** Chromatographic retention times of select peptides from the discovery experiment. Plot shows the consistency of retention times (RT) of K.GTEAAGAMFLEAIPMSIPPEVK.F (blue rectangle), observed in two fractions from multiple LC-MS runs. K.GTEDFIVESLDASFR.Y (red rectangle), on the other hand, was only identified once. In the absence of additional peptides, these single peptides required further information to reduce false protein assignments **D.** Scatter plot of experimental and predicted RTs of peptides from experimental batch 1. RTs were predicted by training a deep learning RT predictor, DeepRT+. Prediction performance is assessed with R^2^ and Δ*t*_95%_ (red dashed lines). *up* = number of unique peptides trained. **E.** Selection of the final 1731 proteins for downstream differential abundance analysis. Use of DeepRT+ salvaged 495 one-hit proteins that would otherwise be removed from downstream analysis. **F.** Dynamic range of all 1731 proteins, ranked in decreasing order of intensity. Each dot represents the median intensity of all intensity values recorded for a given protein across all samples. Intensity is plotted on the log-scale and spans 6.3 orders of magnitude between the high-abundance classical plasma proteins and the low-abundance signaling proteins. Functional groups are based on Putnam’s classification. Red dots highlight representative members in each functional group. Labels are gene names of the corresponding proteins

### One-hit Proteins and Deep Learning for Confident Protein Identification

To examine this further, we differentiated one-hit proteins identified only in single batches of experiments from those identified consistently across multiple batches. We reasoned that identified one-hit proteins consistently identified in multiple independent analyses are less likely to be false identifications, especially if their consistent identification is based on the same unique peptide. These one-hit proteins warrant additional peptide-centric information for protein inference beyond the sequences of the single peptides. **Figure 2B** displays the number of proteins identified in any given number of collective batches. Of the 3803 total proteins (**figure 2B**, cumulative batch ≤7), 51% (*n* = 1941 proteins) were identified based on a single peptide. While the number of proteins identified based on 2 or more peptides increased with increasing number of collective batches, the number of one-hit proteins remained fairly consistent. In particular for cumulative batches three to seven, we found 1698 one-hit proteins that were present in all of them.

To enhance the confidence in the identity of these one-hit proteins and minimize false positive identifications, we employed chromatographic retention time (RT) as additional peptide-centric information and orthogonal to their identification by tandem mass spectrometry. Here, we considered a peptide as confidently identified if, in addition to being a high-scoring peptide by PSM, the observed RT also falls within the RT window expected for that peptide and its corresponding experimental batch conditions. For example, K.GTEAAGAMFLEAIPMSIPPEVK.F, a unique peptide from alpha-1-antitrypsin, A1AT_HUMAN (**figure 2C, supplemental figure 1**, blue rectangles) shows consistent RTs, regardless of the experimental batch or sample fraction the peptide was detected. On the other hand, K.GTEDFIVESLDASFR.Y (**figure 2C**, **supplemental figure 1**, red rectangles) is the only peptide-evidence that translocon-associated protein subunit alpha, SSRA_HUMAN – a one-hit protein – was detected in experimental batch 2.

To determine the RT window expected for these single peptides given the LC-MS conditions of their respective experimental batches, we trained a deep learning-based RT predictor, the DeepRT+ [30], using 80% of the RT of consistently identified peptides for a given experimental batch. We tested the prediction accuracy of the DeepRT+ model with the remaining 20% of the training data and subsequently used the final model to predict the RT of one-hit proteins. We assessed performance of the RT prediction using the coefficient of determination, R^2^, and Δt95%, the minimum time window containing deviations between the observed and the predicted RT for 95% of the peptides (**Figure 2D** and **Supplemental Figure 2**). We found the RT of 495 unique one-hit peptides fell within the Δt95% metric (**Table 2**) and were thus included to a final total of 1731 proteins used for downstream analysis (**Figure 2E**). The dynamic range of all proteins spans 6.3 orders of magnitude and confirms signal from a wide range of abundances in the plasma proteome (**Figure 2F**).

**Table 2.** Summary of DeepRT+ training parameters and results of prediction assessment. Training. Given that each batch of sample has unique LC-MS experimental conditions that uniquely impact RT, seven different models were built for each of the seven batches of experiments. Abbreviations: RT (min): minimum RT for the batch; RT (max): maximum RT; aa: amino acid; up (training): number of unique peptides trained; up (predicted): number of unique peptides whose RTs were predicted; R^2^: coefficient of determination = correlation coefficient for bivariate analysis; Δt95%: deviations between observed and predicted RT that contains 95% of peptides for a given batch of experiment.

**Table 3.** Discovery proteomics data

**Table 4.** Condensin subunit (NCAPH2) linearity assessment

**Table 5.** List of target peptides for PRM and instrument parameters

### Protein Feature Selection and Differential Abundance Analyses

To determine the subset of these 1731 proteins that are most important in discriminating plasma profiles of cases and from non-cases and between baseline and post-operative timepoints, we employed an elastic net regularized regression approach[31]. We found 47 and 64 proteins as signatures of surgical exposure and of delirium, respectively. Principal component analysis (PCA) of study subjects using the subset of protein features demonstrates that delirium cases cluster separately, with marginal overlap between non-delirium controls and baseline samples **(Figure 3A)**. Additionally, plasma profiles of cases and non-cases are clearly separable post-operatively, although they were indistinguishable at baseline (**Figure 3B**). This strongly suggests a temporal relationship between post-operative changes in proteomic signatures and subjects’ surgical exposure and/or related intra-operative physiological events.

**Figure 3:**
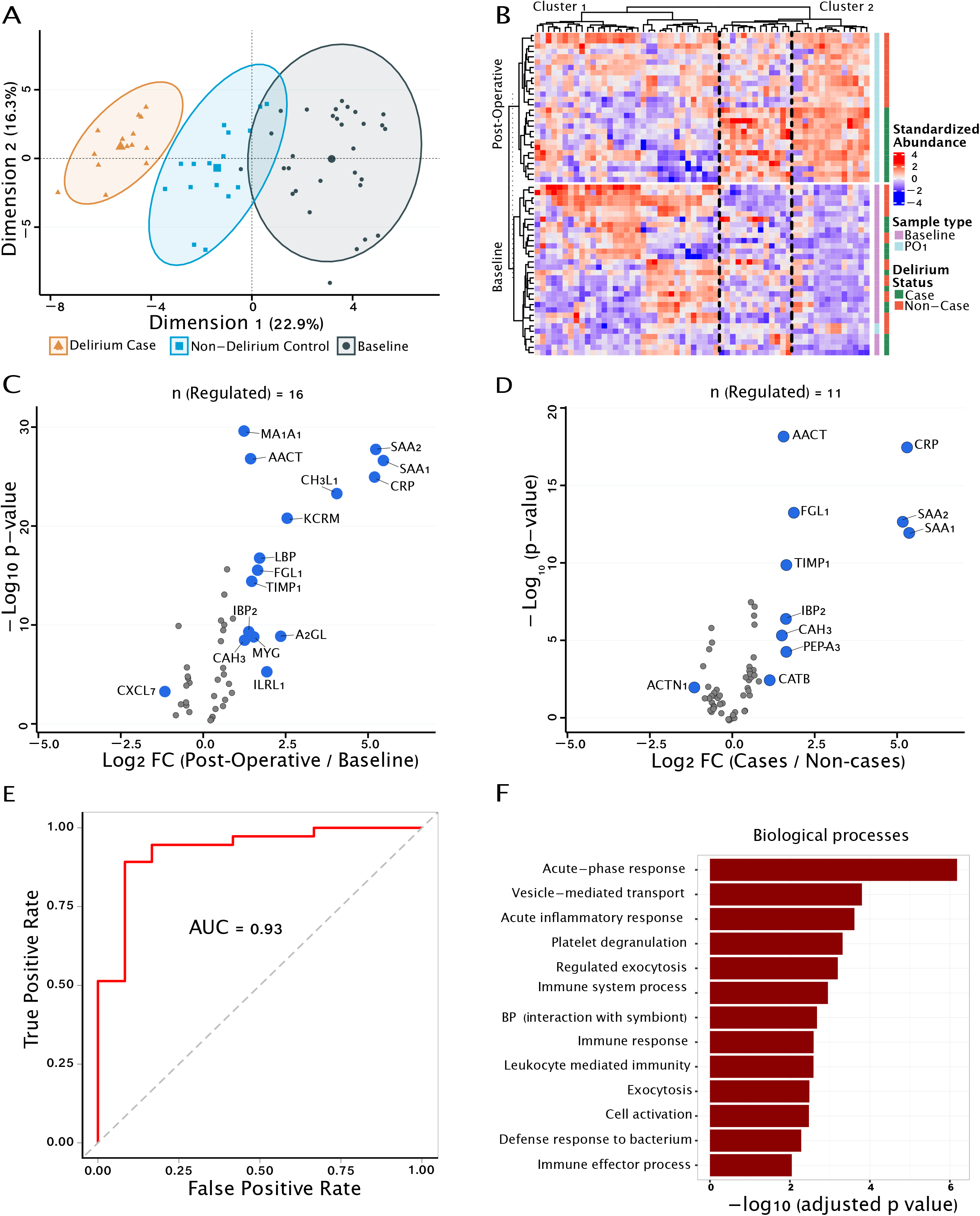
**A.** Principal component analysis of all discovery samples (including replicates). Clustering is based on a subset of 64 proteins identified by the penalized regression approach (ElasticNet) for feature selection. **B.** Hierarchically clustered heatmap of proteomic signatures of delirium cases and non-delirium controls at two time points (baseline and post-operative day 1, PO1). Post-operatively, a subset of proteins (protein cluster 2, dashed lines) shows a higher expression in cases relative to non-cases, although the expression of this subset of proteins was very similar between the two groups at baseline C. Volcano plot of *p*-value (log_10_ scale) vs fold-change (log_2_ scale) of the 47 proteins that explain most of the variation in proteomic profiles of the baseline and post-operative day 1 samples. Blue dot means protein is significantly different at PO1 relative to baseline by at least 2 folds (*p*-value cut-off = 0.05) **D.** Volcano plot of the 64 proteins that explain most of the variation in proteomic profiles between delirium cases and non-delirium controls. **E.** Diagnostic accuracy of the panel of 11 differentially abundant proteins that discriminate cases from non-cases. **F.** Functional analysis of biomarkers for biological processes enriched among the panel of 11 differentially abundant proteins that discriminate cases from non-cases.

Furthermore, we quantified the extent of changes in biomarker levels before and after surgery (**Figure 3C**) and between cases and non-cases (**Figure 3D**). When using the proteins identified as a signature of delirium (**Figure 3D**), we observed a diagnostic accuracy of 93% in discriminating cases from non-cases (**Figure 3E**). Functional analysis of the biomarker panel for biological processes shows acute inflammatory response and activation of the immune system as the most significantly enriched functional pathways, predominantly in the extracellular region (**Figure 3F** and **Supplemental Figure 3**).

### Biomarker Verification

For further evaluation of peri-operative proteomic differences between cases and non-cases, an independent set of plasma samples was used to verify biomarkers discovered *a priori* (**Figure 4)**. Here, we used parallel reaction monitoring (PRM) as the targeted approach and employed label-free quantification (LFQ) as orthogonal methods different from the TMT approach used in the discovery phase. To ascertain the degree to which changes in protein concentration in the complex background of plasma are quantifiable, we artificially modified six biological replicates of a pooled plasma sample with the addition of exogenous proteins: (1) equal amounts of a non-human (*Schizosaccharomyces pombe*) homolog of the serine/threonine-protein kinase Chk2 (CDS1 in *S. pombe*); and (2) increasing concentrations of heavy-labeled AQUA peptides[32, 33] of human condensin-2 complex subunit H2 (CNDH2). From this experiment, we estimate a limit of quantification of ∼1fmol on column (**Figure 5A**), with negligible impact on target protein quantification due to matrix effects from large (16-fold) variations in the concentration of a non-target protein in the matrix (**Supplemental Figure 4**).

**Figure 4:**
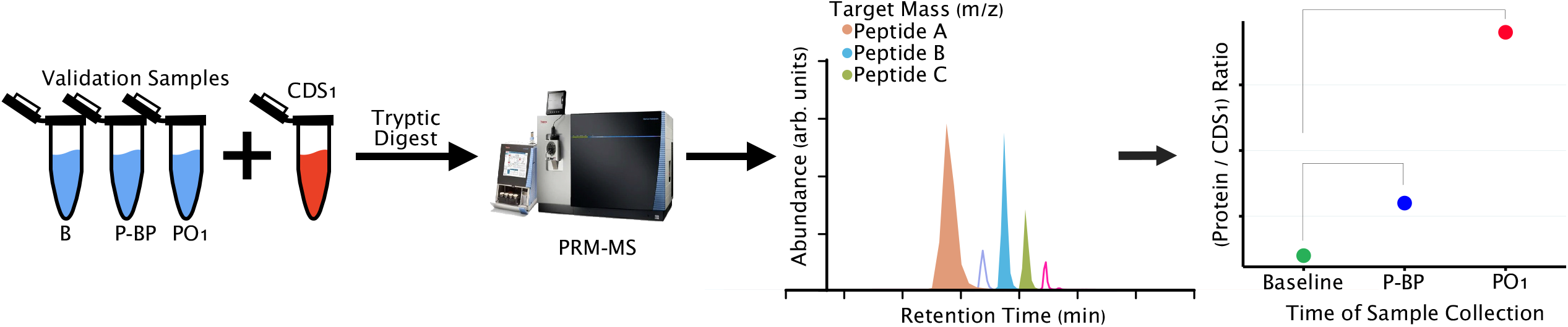
Biomarker Validation. Validation samples included baseline (*B*), post-bypass (*P-BP*) and post-operative day 1 (*PO1*) samples. To each unlabeled validation sample, an equimolar amount of CDS1, a protein from *S. pombe*with no sequence overlap to human proteins previously expressed and purified from bacteria, was added as a reference standard to control for run-to-run variations. Select tryptic peptides of regulated proteins from the discovery phase were targeted for quantification using via PRM-MS. Concentrations of each biomarker were analyzed for changes across the sampling time points (*B*, *PB*, *PO1*). Hypothetical data are depicted as exemplars.

**Figure 5:**
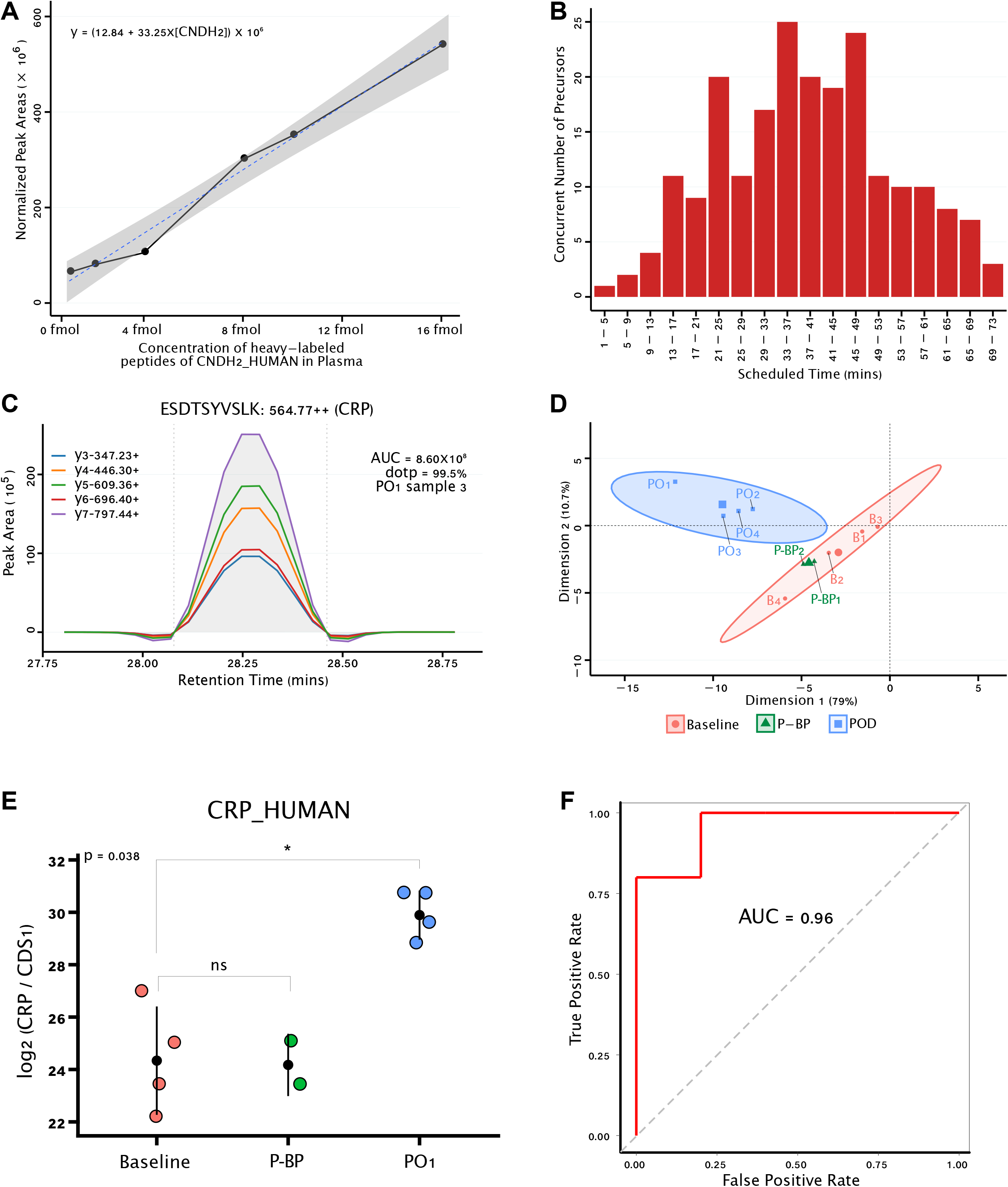
**A.** Normalized peak areas of CNDH2_HUMAN condensin subunit with increasing concentrations of its heavy-labeled stable isotope standards spiked into a background matrix of plasma. Grey area is the 95% confidence band of the regression line of fit: y = (12.84 + 33.25x[CNDH2]) x 10^6^ **B.** Number of precursors monitored concurrently during five-minute windows across the 78-minute gradient for used for validation experiments. **C.** Representative extracted ion chromatogram (XIC): the five most intense fragment ions of the CRP_HUMAN peptide ESDTSYVSLK, co-eluting at 28.3mins. All other peptides were quantified similarly with a minimum of five transitions consistent across all samples, a minimum dot product (dotp) of 95% and manual inspection for distinct peak boundaries and interference-free transitions. **D.** Principal component analysis of all validation samples. Notable here is the clustering of post-bypass samples together with the baseline, signaling similar proteomic signatures between the two timepoints. **E.** Representative plot of differential abundance analysis of validated proteins for the candidate biomarker C-reactive protein (CRP), showing changes across the three sample collection time points: baseline, post-bypass (P-BP) and post-operative day one (PO1) **F.** ROC analysis of the discriminatory power of the validated panel of biomarkers

For candidate biomarker verification, we developed parallel reaction monitoring (PRM) methods through an iterative optimization process (**Supplemental Figure 5**). We monitored 153 unique peptide sequences (212 total precursor ions including the observed range of charge states) from the union of 18 differentially abundant proteins as PRMs that were distributed across the entire LC-PRM elution gradient (**Figure 5B)**. For example, we monitored the abundance of the peptide ESDTSYVSLK from C-reactive protein as a doubly-charged ion via five individual y-ions in our PRM method via Skyline (**Figure 5C**) in each verification sample. The PRM methods we employed required the following minimum criteria for peptide quantification: a consistent minimum of 5 transitions in all samples, a minimum dot-product of 95% and manual inspection of all peaks for interference-free co-eluting transitions with distinct peak boundaries. 65 precursors from 13 proteins met these criteria for downstream analysis (**Supplemental Table 3**). Unsupervised clustering based on the quantification of these candidate biomarkers shows that post-operative samples aggregate separately from post-bypass and baseline samples (**Figure 5D**). This is further confirmed by statistical comparison of biomarker levels between the sampling timepoints (**Figure 5E** and **Supplemental Figure 6**).

Seven biomarkers (A2GL, AACT, CH3L1, CRP, LBP, MA1A1 and SAA1/SAA2) were significantly increased at post-operative day one (PO1) relative to baseline in this validation cohort. Four razor peptides were shared between SAA1 and SAA2. However, no peptides unique to either SAA1 or SAA2 met the minimum quantification criteria for PRM verification. Similarly, none of the precursor peptides of CAH3, EFNA1, FGL1 or PEPA4 met PRM quantification criteria. Regardless of statistical significance, we observe that these candidate biomarker levels show a consistent increase in abundance between baseline and PO1 (**Supplemental Table 3**). This panel of differentially abundant candidate biomarkers yields a discriminatory power of 96% (84.9 – 100%) between cases and non-cases (**Figure 5F**).

## Discussion

This unbiased proteomic analysis of samples from a prior nested case-control study is the deepest unbiased plasma proteomic profiling for potential biomarkers of delirium to date. We employed a rectangular biomarker workflow[34] to both discover and verify biomarkers of post-operative delirium on a single mass spectrometry platform without the use of traditional affinity-based verification methods. Dominated by one-hit wonders, our focus on the low-abundance proteome presented us with the challenge of protein inference, for which we applied deep learning to recover pertinent orthogonal peptide chemical information and salvage a significant number of these one-hit proteins. We identified 3808 proteins by isobaric quantitative multiplexed proteomics, 16 of which were differentially abundant post-operatively from baseline levels, and 11 of which were differentially abundant in cases relative to controls. This includes proteins with well-documented associations with delirium, such as CRP, CH3L1, AACT, TIMP1, as well as new ones not previously associated with delirium, including SAA, CATB and PEPA3. Using an independent set of samples, we attempted to verify the union of these candidate biomarkers and found a 96% accuracy in correctly identifying delirium patients for those for which quantification was possible. Collectively, our findings show a temporal association between intra-operative events (i.e., surgical insult, administered anesthesia, etc.) and proteomic changes associated with phenotypic delirium.

The prevailing mechanistic hypothesis of delirium is one of acute neurocognitive disruption triggered by system-wide inflammation[18, 19]. In our study, functional analysis of the post-operatively dysregulated biomarkers suggests a system-wide activation of the inflammatory cascade and related immunological reactions. Data on the associations between delirium and acute-phase reactants (APR) such as CRP is ubiquitous[20, 35–38]. Although known APRs correlate well with the severity of inflammation, their usefulness as biomarkers is limited as they are not specific to delirium. We, however, found additional acute-phase reactants that may shed a new light on delirium.

Human serum amyloid A (SAA) is a collective name for a group of polymorphic proteins functionally associated with high-density lipoprotein (HDL). By the regulation of their synthesis, they are grouped into the acute phase isotypes (a-SAA: SAA1, SAA2 and SAA3) and the constitutive isotype (c-SAA: SAA4)[39, 40]. Although predominantly secreted by the liver, extra-hepatic production occurs in the brain and may be more relevant in neurocognitive disorders such as Alzheimer’s disease[41–44]. SAA has cytokine-like effects which likely provokes blood brain barrier (BBB) dysfunction, induces depressive-like behavior in mice and may impair cognition in human subjects[45–48]. In the present study, we found SAA1 and SAA2 were both upregulated post-operatively in delirium cases by over 5 folds (*p* value < 0.001). This is the first mention of SAA in the context of delirium and warrants further studies to formally credential this association with the condition.

The cysteine protease cathepsin B (CATB) has previously been quantified as an AD-related biomarker and correlates with mini-mental state examination (MMSE) scores [49–52], but its association with delirium is unknown. It is an inflammasome that promotes IL-1beta maturation and secretion[52]. It also has a beta-secretase activity, capable of cleaving amyloid precursor protein into amyloid beta [53]. Given that cases and non-cases in our study were matched by baseline neurocognition and tMOCA scores were statistically controlled for, upregulation of CATB in delirium cases may indicate a common pathophysiological starting point in the continuum of neurocognitive disorders, of which delirium and AD are a part. Generally recognized as the first enzyme to be discovered, pepsin (PEP-A) is the native acid protease of the stomach[54]. Blood pepsin is an established biomarker of gastric mucosal integrity, and plasma levels correlate with the degree of mucosal damage[55–58]. Cardiac surgery and CPB places enormous physiological stress on the body. Through the cholinergic anti-inflammatory reflex, the body attempts to ameliorate the stress by increasing vagal tone[59–62] which manifests as gastric acid production. Normally, small amounts of secreted pepsin (∼1%) may be found in blood and urine[63], but with increased acid production, this proportion may be higher. In the discovery phase of our study, differentially abundant PEP-A levels in cases relative to non-cases (1.64-fold increase, *p* value < 0.001) despite pre-operative proton-pump inhibitor administration in the study subjects suggests a peculiar association between plasma PEP-A levels and delirium. At present, we are unable to explain the relationship, if any, between increased vagal tone and neuroinflammation.

The independent association between CPB and delirium remains an ongoing debate and data on the relationship is conflicting. On the one hand, the use and duration of extracorporeal circulation is reported to increase the risk of delirium[64–66]. Some authors, on the other hand, have reported no associations between delirium incidence and CPD duration[67, 68]. In our cohort, there was no statistically significant difference in aortic cross-clamp time or duration of bypass between delirium cases and non-cases.[28] To determine the impact of CPB in our cohort, we compared post-operative plasma profiles to baseline regardless of the case/non-case status of subjects. We found 16 dysregulated proteins, most of which have been characterized as non-specific markers of surgical exposure[69–71]. A striking observation in our study is the similarities in proteomic signatures between cases and non-cases at baseline, despite a clear difference at post-operative day one. Previous studies have shown that post-operative delirium cases are likely to be in a heightened pre-operative inflammatory state [20, 37, 60, 72–75], which makes them more vulnerable to intraoperative stressors. In our study, similarities in the levels of identified biomarkers at baseline suggests otherwise.

The main strength of the present study is in its unbiased, hypothesis-generating approach to identify potential biomarkers of delirium. This lays the groundwork for future studies and highlights new frontiers in delirium research yet to be explored. Translational utility from the research bench to the patients’ bedside requires that the biomarker readout in the discovery phase is independent of the measurement approach used for their discovery[76]. For this reason, we validated discovered biomarkers using label-free quantification, which is orthogonal to the TMT-based measurements in the discovery phase of our study. Our choice of PRM-MS over traditional affinity methods for validation (e.g., ELISA) is further premised on the fact that affinity methods are semi-quantitative with inter-operator variability in quantification, have limited dynamic range and require larger amounts of sample. In addition to the requirement for peptide antigenicity, antibody cross-reactivity limits multiplexing (i.e., how many proteins can be validated at a time)[77]. All proteins needing validation require antibodies, a step that takes considerable amount of time to develop and can be cost-prohibitive if commercial options are not available[78]. This, in fact, is a long-standing bottleneck in clinical biomarker workflow[79].

Our study is, however, not without limitations. First, sample sizes for both the discovery and validation phases may have limited statistical power in detecting differences in the levels of many other biomarkers. In our cohort, the CAM test was administered daily after surgery. In our statistical analysis, we did not correct for the effects of retesting on repeated test administration in this cohort. In the discovery phase, our interest in the low-abundance plasma proteome required an immunodepletion step to remove the majority of the top 14 most abundant plasma proteins. The extent to which this experimental step contributed to the removal of other proteins through their specific or non-specific binding was not ascertained. Although isotypes SAA1 and SAA2 each had unique peptides in the discovery phase, only the razor peptides met the criteria for quantification in the validation phase and were thus undistinguishable. Similarly, peptides from CAH3, EFNA1 and PEPA3 did not meet the minimum quantification criteria for verification by PRM, and peptides from FGL1 were not detected at all in any of the verification samples by PRM.

In summary, diagnostic biomarkers of delirium are urgently needed for accurate case identification, long-term risk stratification and for molecular characterization of delirium. In this study, we discovered a panel of biomarkers through the unbiased comparative analyses of baseline and post-operative plasma samples of delirium cases and non-cases. We underscored the importance of brain-specific biomarkers such as SAA and CATB and their possible role in the pathophysiology of delirium. In the long-term, it is in our research interests to rigorously test their associations with delirium and ascertain how these biomarkers change over time in a larger independent cohort.

## Data Availability

All data produced in the present work are contained in the manuscript

## Abbreviations

CABG: coronary artery bypass grafting
CAM: confusion assessment method
CPB: cardiopulmonary bypass
POD: post-operative delirium
SPS-MS: synchronous precursor selection – mass spectrometry
TMT: tandem mass tagging

## Acknowledgments

Funding support for this work was supplied by the Burroughs Wellcome training grant to KW, the National Institutes of Health (R01 GM122846) to SAG, and K08 GM134220 and R03 AG060179 to SS. The heavy-labeled peptides of CNDH2 condensin subunit were also provided by Dr Giovanni Bosco (Dartmouth College).

## Materials and Methods

### Study Design and Patient Enrollment

Subjects in this nested case-control study were selected from the parent study, a randomized double-blind trial conducted on subjects who underwent coronary artery bypass grafting (CABG) with cardiopulmonary bypass (CPB) between July 2015 and July 2017 at the Beth Israel Deaconess Medical Center (BIDMC) in Boston MA. The trial was registered with ClinicalTrials.gov (NCT02591589, https://clinicaltrials. gov/ct2/show/NCT02591589, principal investigator: Shahzad Shaefi, registration date: October 29, 2015). Institutional review board (IRB) approval 2014P000398/33 was amended for the purposes of this current study on 09/17/2021 by the Committee on Clinical Investigations at the BIDMC. Details of enrollment, subject randomization and treatment allocation in the parent study are published elsewhere [28, 80]. Briefly, patients aged 65 years or older who were booked for elective CABG requiring CPB were eligible. The primary objective was to examine the temporal relationship between intra-operative oxygen treatment and post-operative neurocognitive function as measured by the telephone-based Montreal Cognitive Assessment (tMOCA) score. Patients were assessed for delirium as a secondary endpoint using the confusion assessment method (CAM). Patients were excluded if they were undergoing emergent CABG, if they required single-lung ventilation, CABG without CPB, intraoperative balloon counter-pulsation or mechanical circulatory support. All patients provide informed consent. 15 subjects were randomly selected for proteomic profiling in this nested case-control study. Because quantitative studies on the effect size of delirium biomarkers using mass spectrometry is largely unexplored, formal power analysis was not done.

### Sample Collection

Whole blood samples at baseline, post-bypass (P-BP) and on post-operative day one (PO1) were collected into 4mL EDTA-treated tubes (BD Diagnostics) and centrifuged immediately at 200*g* at room temperature for 10 min. Resulting plasma was stored at - 80°C until they were thawed for aliquots used here for proteomic profiling.

### Chemicals and Reagents

All LC-grade chemicals are marked with asterisk (*): Dithiothreitol (DTT), 4-(2-hydroxyethyl)-1-piperazineethanesulfonic acid (EPPS), Tris (hydroxymethyl) aminomethane (Tris), formic acid* and acetonitrile* were purchased from Sigma-Aldrich. Methanol* was obtained from Fisher. Trypsin Protease, SDS, 2-iodoacetamide (IAA), High Select Top14 Abundant Protein Depletion Mini Spin Columns and TMT 11 plex kit were acquired from Thermo Fisher Scientific.

### Sample Preparation analysis

#### Sample Immunodepletion

Buffer exchange on single-use High Select Top14 Abundant Protein Depletion mini-spin columns (ThermoFisher Scientific) was performed twice using 200 µL of 50mM Tris [pH 8.1] / 50mM NaCl. 10 µL of each plasma sample was applied to the mini-columns, incubated at −4°C with gentle end-over-end mixing for 15 min, according to manufacturer’s instructions. Flowthrough were collected by centrifugation at 1000*g* for 2 min into 2mL Eppendorf tubes. Concentrations of the depleted samples were obtained using the Pierce BCA Protein Assay Kit (Thermo Scientific) at 562 nm absorbance per manufacturer’s instructions.

### Digestion and Labelling for Biomarker Discovery

Depleted samples were treated with SDS (2% final) and DTT (2mM final) for denaturing at 75°C for 15 min. Samples were cooled to room temperature before alkylation with IAA (7mM final) at room temperature in darkness for 30 min and quenched with DTT (additional 2mM final) for 10 minutes. Proteins were isolated by single-pot solid-phase-enhanced sample preparation (SP3) and digested to peptides in EPPS buffer overnight at 30°C with 1:50 w/w trypsin (Promega^TM^). Tryptic peptides were labeled with TMT-11 plex reagent for 1 hr according to manufacturer’s instructions. Two channels in each set of TMT-11 plex were reserved for pooled plasma to be used as bridge samples for technical control. Labeling efficiency of at least 95% was confirmed on a 1-hr gradient before pooling. Labeled tryptic peptides were then desalted on an OASIS µHLB (Waters) and subsequently dried by vacuum centrifugation prior to off-line HPLC fractionation on a pentafluorophenyl (PFP) column as described previously [29]. 48 fractions were concatenated into 12 fractions for LC-MS/MS analysis. All samples were prepared in duplicates.

### Digestion for Biomarker Validation

Equal amounts of recombinant purified CDS1 protein were added to each depleted sample before treatment with SDS (2% final) and DTT (2mM final) for denaturation and alkylation as described above. Proteins were isolated by single-pot solid-phase-enhanced sample preparation (SP3) and digested to peptides in 50mM ammonium bicarbonate buffer overnight at 30°C with 1:50 w/w trypsin (Promega^TM^). In a separate experiment to check for signal linearity, increasing concentrations of heavy-labeled peptides of CNDH2 condensin subunit were added to the samples at this point. Tryptic peptides were desalted on an OASIS µHLB (Waters) and dried by vacuum centrifugation. All samples were run in duplicates.

### LC-MS/MS

All data were acquired on an Orbitrap Fusion Lumos Tribrid instrument (ThermoFisher Scientific, San Jose, CA) equipped with EASY-nanoLC 1200 ultra-high pressure liquid chromatograph (ThermoFisher Scientific, Waltham, MA). Dried peptides were resuspended in 5% methanol / 1.5% formic acid and injected onto a 35-cm long / 100-µm (inner diameter) in-house pulled analytical column packed with Reprosil C18 stationary phase particles. Discovery samples were separated on 120-minute gradient, and validation samples on a 60-min gradient, at 350nL/min flow rate. Acquisition parameters included 120,000-resolution at MS1, AGC target value of 5.0×10^5^, scan range of 350 – 1250 m/z and maximum injection time of 100ms. For the TMT-labeled peptides, the top eight MS2 peaks were selected for further fragmentation at 55% normalized high-collision energy (HCD) via SPS-MS3 for quantification of reporter ions in the scan range of 110 – 500 m/z. For the label-free peptides in the validation phase, MS2 scans were generated at 30,000 resolution and AGC value of 2.5×10^5^, using 30% normalized collision energy (HCD).

### Bioinformatics

#### Peptide Spectral Matching

Acquired data (in .raw format) were searched using COMET [81] against a target-decoy version of the human proteome (Uniprot, downloaded in 2020 and 2022, for the discovery and validation phases respectively). The fasta for the validation phase was appended with sequences from CDS1_SCHPO. Search parameters included a mass tolerance of 20ppm, maximum missed cleavages of 3, carbamidomethylation of cysteine as fixed modification and oxidized methionine as variable modification. In addition, the mass of 229.162932 Da was added to the N-termini and lysine residues of all peptides as fixed modification for the TMT data. A false discovery rate (FDR) of 1% was applied at the peptide level and final list of PSMs were filtered using XCorr and delta XCorr. All data were subsequently imported into R environment for statistical computing (v4.1.1) and Python programing language (v3.8) for downstream analyses [82, 83].

### TMT Data Wrangling and Normalization (Discovery Phase)

After correcting for differential sample loading, the ratios of sample proteins to their respective bridge proteins were computed. Here, data from bridge samples was used for quality control and to correct for batch-to-batch technical variations. Values were subsequently log-transformed and mean-centered. Data from all batches were combined and analyzed for possible outlier observations using OutlierDM R Package. Proteins were removed if their frequency of observation was less than half of all samples. For one-hit wonders in each batch of experiment, a retention time (RT) predicting model was built in Python using DeepRT+ as described by Ma, Ren [30]. Prediction performance was assessed with coefficient of determination (R^2^) and delta-t95% (Δ*t*_95%_). Δ*t*_95%_ is the minimum time window containing deviations between the observed and the predicted RT for 95% of the peptides. Peptides with RT outside the Δ*t*_95%_ range were excluded from downstream analysis. Missing entries in the data were imputed by making random draws from the left tail of the gaussian distribution of the entire log-transformed data matrix (using −2.5 SDs from the mean, width = 0.3).

### Protein Feature Selection and Differential Abundance Analyses

To determine the subset of protein features that differentiated cases from non-cases, or postoperative expression profiles from baseline, Elastic Net algorithm was used [31]. This is a regularization and feature selection method with good performance on high-dimensional data (i.e., an *n*×*p* data with very large *p* proteins but small *n* samples). Elastic Net is insensitive to features that dominate the matrix (e.g., albumin) and likely suppress signal from low abundance predictors and skew model coefficients. In addition, Elastic Net is a good choice if overfitting and multicollinearity (or protein features that are highly correlated and essentially communicate the same information) are a concern. Tuning parameters were achieved by grid optimization with a five-fold nested cross-validation where the last fold was held out for testing. The average of hyperparameters from all folds were computed and used to build the final model.

Using the subset of protein features, an unsupervised visualization of the data was achieved with principal component analysis (PCA). Hierarchical clustering was employed to check for reproducibility of replicate samples and inherent sample clusters, and together with a heatmap, the overall protein expression patterns. Here, clustering was achieved using Ward’s clustering algorithm.[84] Briefly, Ward’s minimum variance method begins with singleton clusters and recursively merges them by minimizing the total within-cluster variance as the objective function. After this point, protein values for any given biological replicates were summarized as means prior to differential abundance analyses. Two-way comparison for differential abundance was achieved by Student’s *t*-test, assuming unequal variance. Differential abundance analysis was visualized with volcano plots. Because statistical comparison was done for only a subset of proteins, no correction for multiple hypothesis testing was done. Proteins were deemed differentially regulated between conditions if there was a statistically significant *t*-test (*p* value cutoff ≤ 1.5) and a log2 fold-change of at least ±1. This fold-change cutoff was selected to prioritize a panel of biomarkers with significant changes between conditions that is unlikely to be due to chance.

### PRM Label-free Data Procession (Validation Phase)

Raw files were imported into Skyline v21.2.0.369 [85]. Precursor peptides with modifications other than carbamidomethylation of cysteine (as fixed modification) or oxidized methionine (as variable modification) were excluded. Peptide quantification criteria was defined as follows: (1) consistently identified precursors across all validation samples, (2) with maximum of two missed cleavages, (3) a consistent minimum of five transitions, and (4) at least 0.95 dop-product with the spectral library of chromatograms. All peak boundaries were manually inspected for interference-free co-eluting transitions before peak areas were integrated at the MS2 level. For any given precursor peptide, the five most intense fragment ions in the *m/z* range of 120 – 1500 were used for quantification. Final dataset was exported as .csv and analyzed in R environment for statistical computing (v4.1.2; R Core Team 2021). No imputations were required in the validation data. Data was normalized by computing peak area ratios relative to CDS1_SCHPO to correct for run-to-run variations. For each protein biomarker, Kruskal Willis global test was first used followed by post-hoc Mann-Whitney U test for pairwise comparisons of the normalized peak areas between the different sample collection timepoints (baseline, post-bypass and post-operative day 1).

### Data accessibility statement

Datasets from the discovery and validation phases are available as supplemental material.

## Supplemental Figures

**Supplemental Figure 1.**
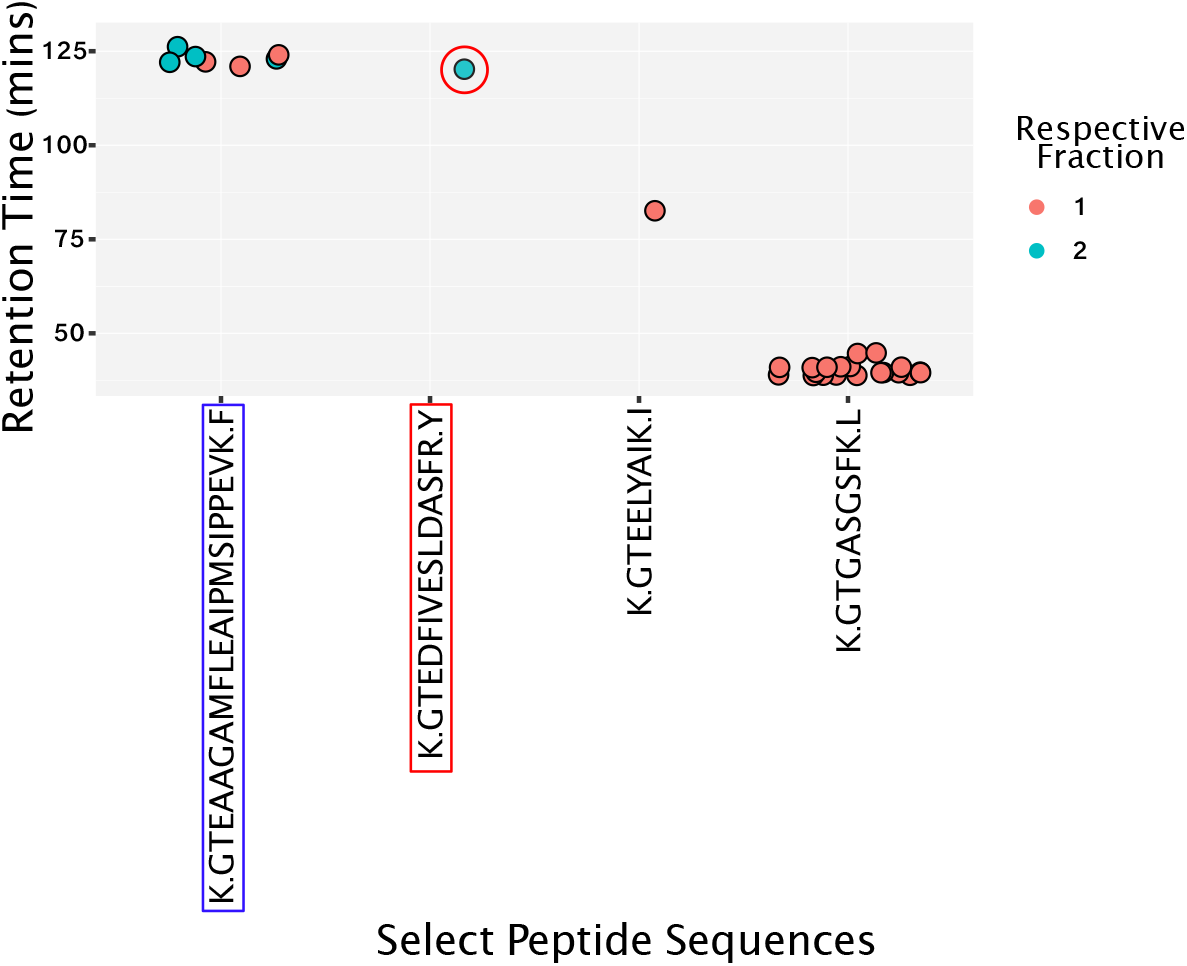
Chromatographic retention times of select peptides, showing consistency of RT and adjacency of sample fractions from which they were identified

**Supplemental Figure 2.**
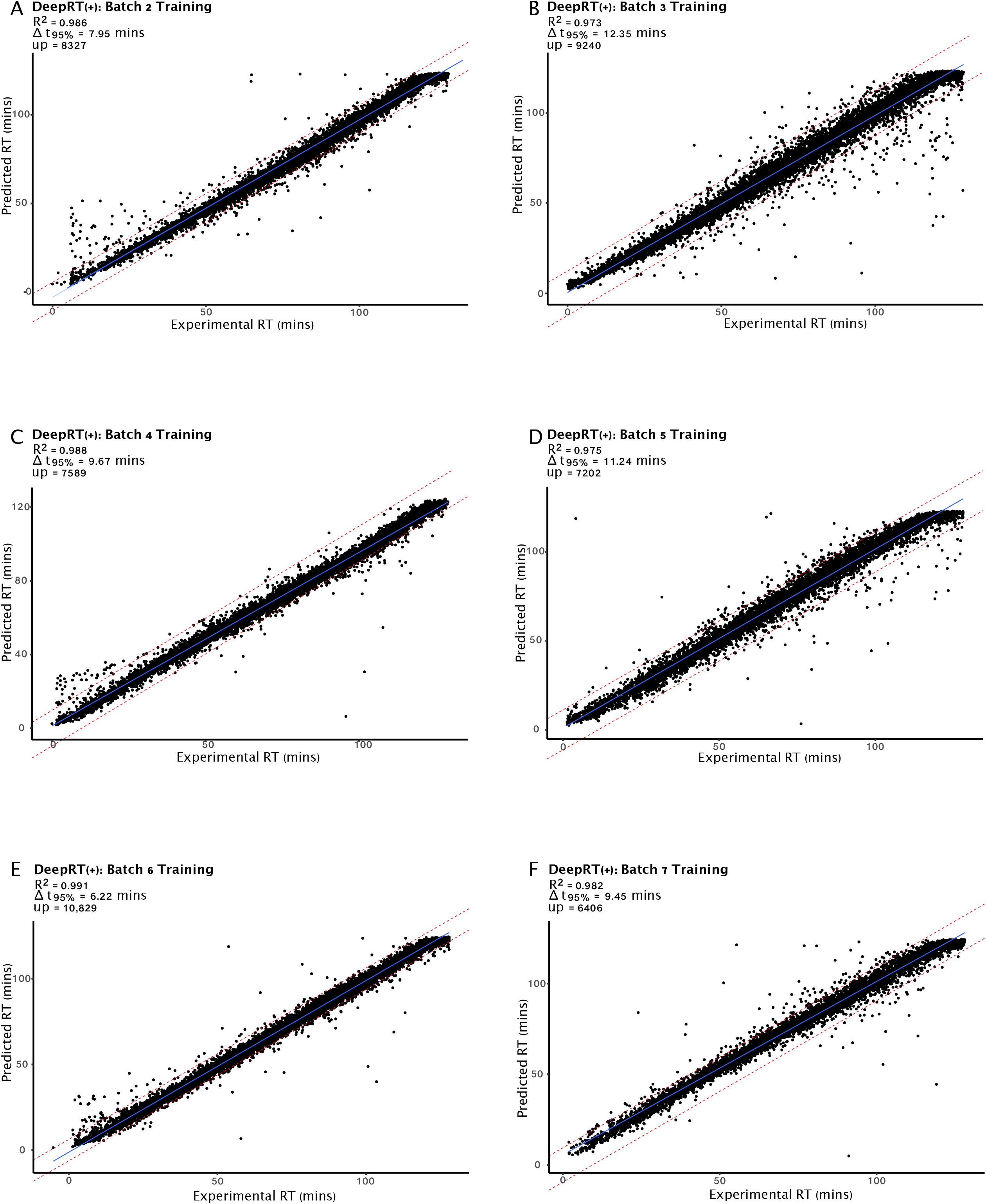
Scatter plot of experimental and predicted RTs of peptides from experimental batch 2 - 7. *up* = number of unique peptides trained

**Supplemental Figure 3.**
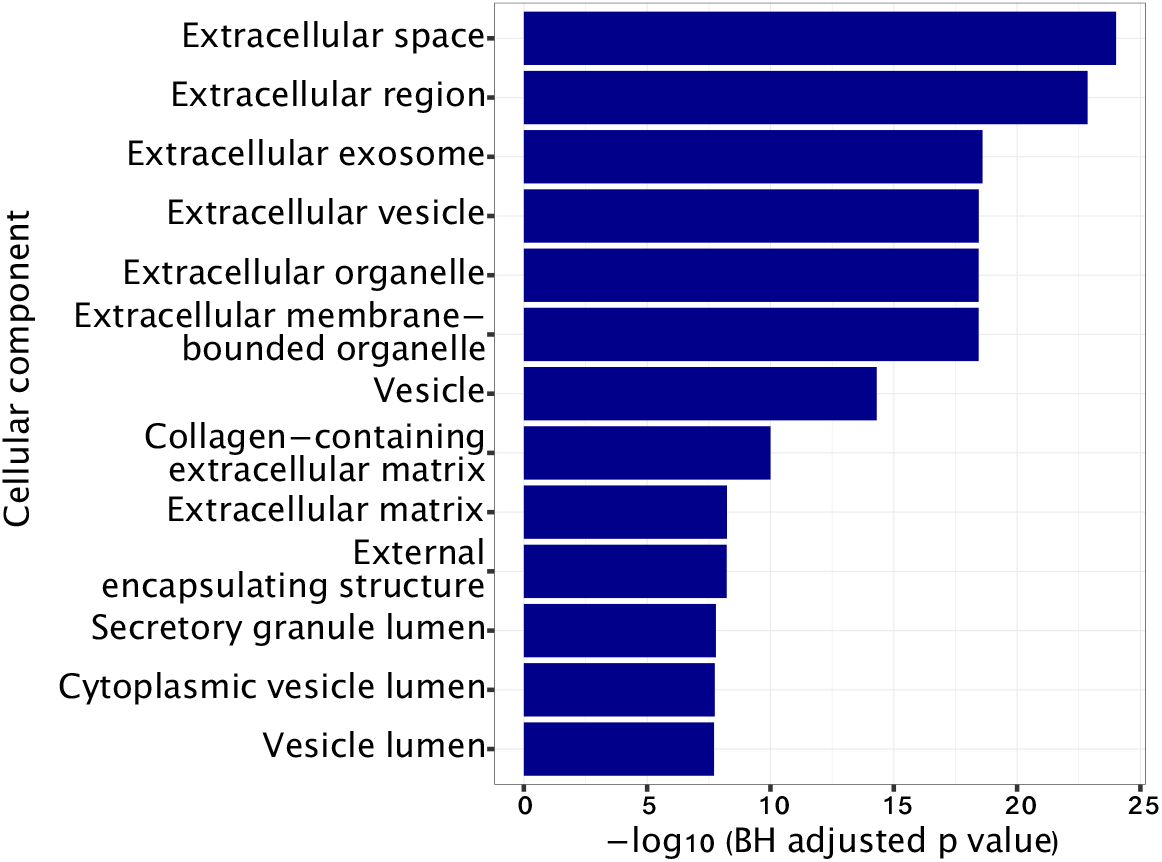
Functional analysis of biomarkers for enriched cellular components

**Supplemental Figure 4.**
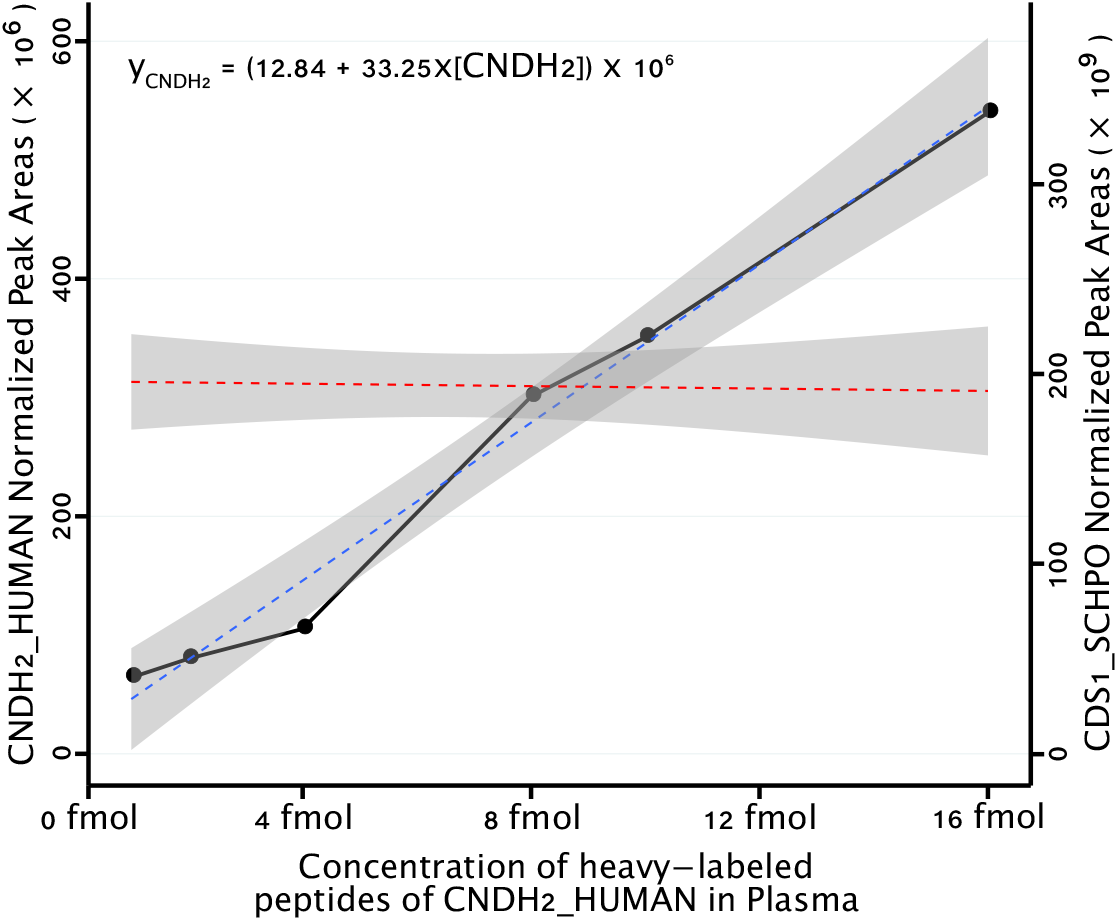
Normalized peak areas of CNDH2_HUMAN condensin subunit, superimposed with CDS1-SCHPO against increasing concentrations of its heavy-labeled stable isotope standards spiked into a background matrix of plasma.

**Supplemental Figure 5.**
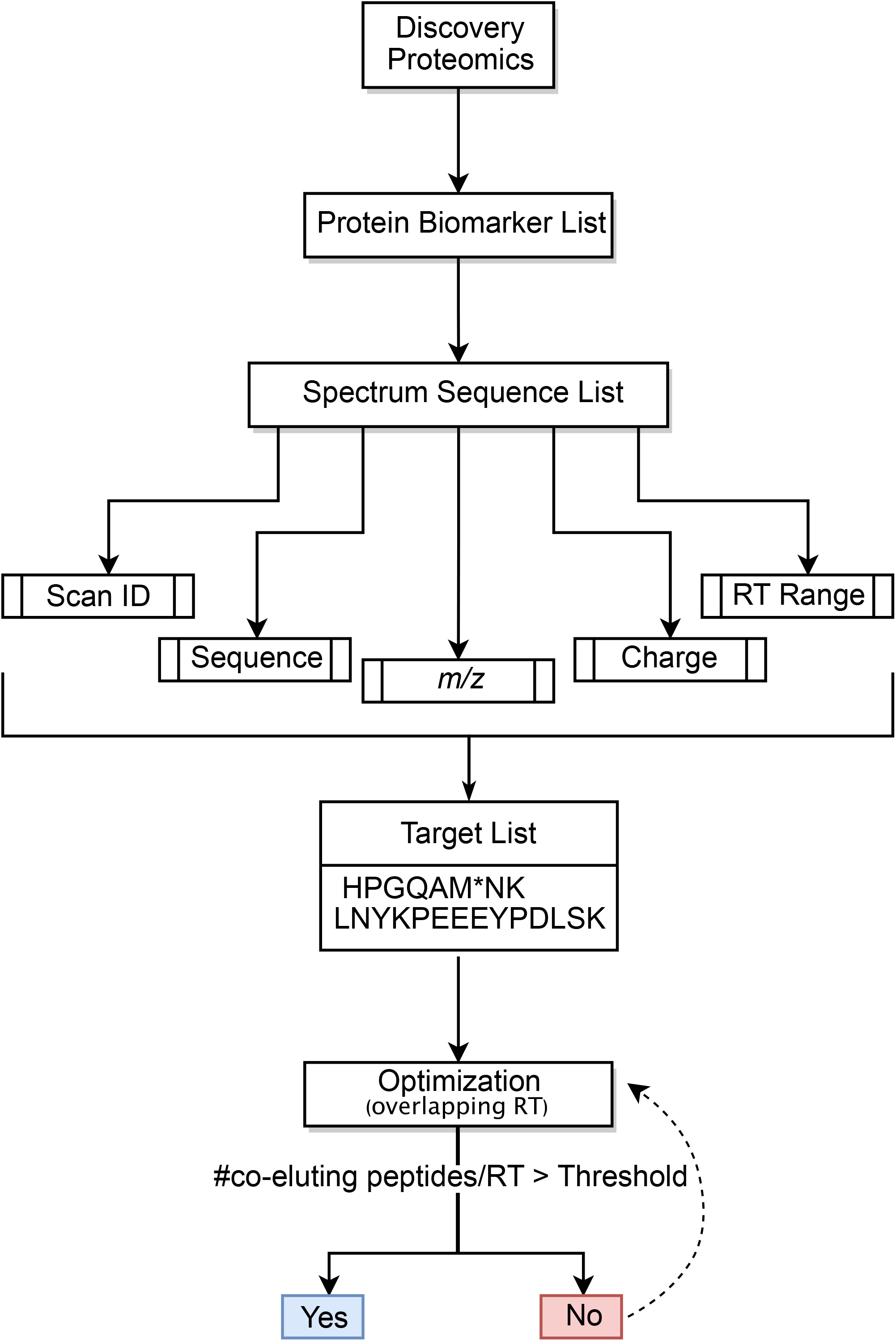
Flowchart of PRM method development

**Supplemental Figure 6.**
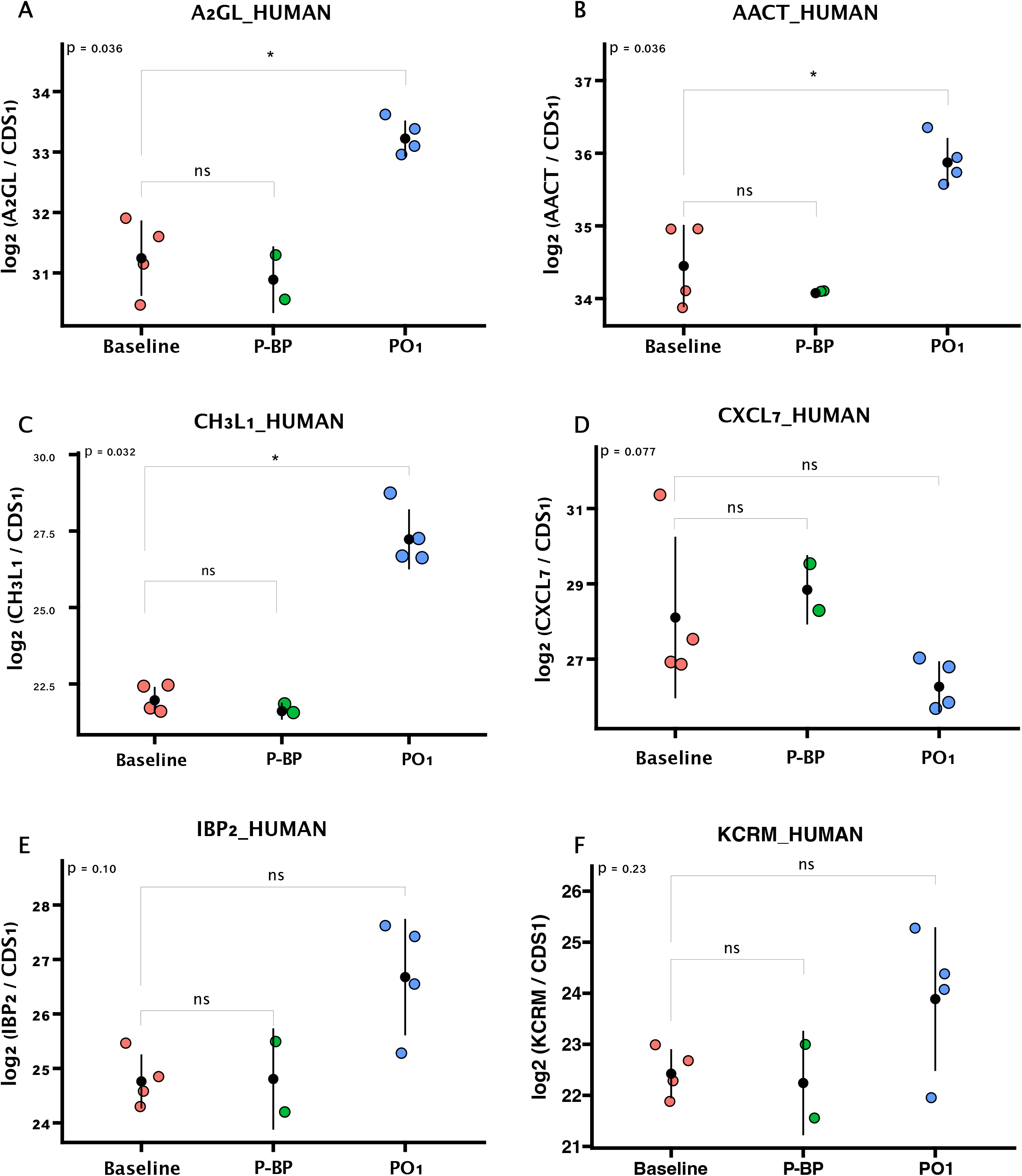
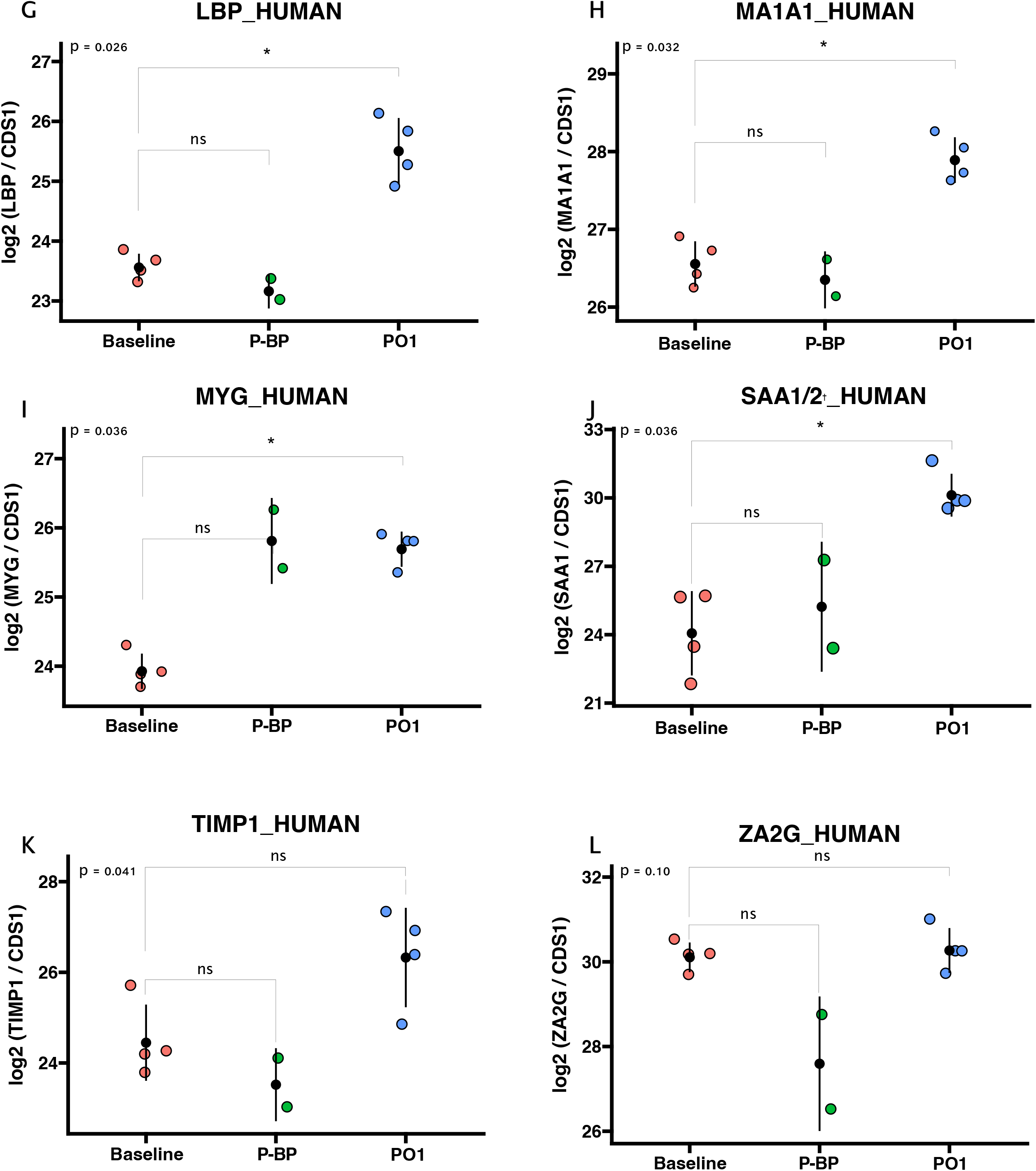
Differential abundance analysis of validated proteins, showing changes across the three sample collection time points: baseline, post-bypass (P-BP) and post-operative day one (PO1). ^†^: SAA1 and SAA2 could not be distinguished in the validation phase as none of the peptides unique to them met the quantification criteria.

## Notes

### Competing Interest Statement

The authors have declared no competing interest.

### Clinical Trial

NCT02591589

### Clinical Protocols

https://clinicaltrials.gov/ct2/show/NCT02591589?term=NCT02591589&draw=2&rank=1

### Author Declarations

Institutional review board (IRB) approval was provided by the Committee on Clinical Investigations at the Beth Israel Deaconess Medical Center (BIDMC) in Boston MA

