## Supplementary material for "Intraoperative Plasma Proteomic Changes in Cardiac Surgery: In Search of Biomarkers of Post-operative Delirium": table_1,2.docx

|  | Delirium Cases | Non-Delirium Controls |
| --- | --- | --- |
| Sample | *n* = 7 | *n* = 8 |
| Age | 70 (±5.0) | 71 (±4.4) |
| Sex (male) | 7 (100%) | 8 (100%) |
| *t*MOCA | 17 (±2.3) | 17 (±1.9) |
| Hyperoxia | 4 (57%) | 4 (50%) |

Table 2:

| Training Parameters | | | | | | Results | | |
| --- | --- | --- | --- | --- | --- | --- | --- | --- |
| Batch | RT (min) | RT (max) | max aa length | up (training) | up (predicted) | $\Delta$t95% | R^2^ | Eliminated |
| 1 | 4.1 | 129.2 | 46 | 8414 | 776 | 6.89 | 0.989 | 145 |
| 2 | 5.2 | 129.4 | 45 | 8327 | 758 | 7.95 | 0.986 | 104 |
| 3 | 10.2 | 129.7 | 44 | 9240 | 859 | 12.35 | 0.973 | 127 |
| 4 | 9.3 | 129.5 | 43 | 7589 | 646 | 9.67 | 0.988 | 121 |
| 5 | 10.5 | 128.7 | 40 | 7207 | 749 | 11.24 | 0.975 | 129 |
| 6 | 4.4 | 128.6 | 43 | 10829 | 849 | 6.22 | 0.991 | 170 |
| 7 | 11.1 | 128.7 | 42 | 6406 | 734 | 9.45 | 0.982 | 120 |
